# The Associations of Cerebral Blood Flow and White Matter Hyperintensities with Tau and Amyloid-beta Across the Alzheimer’s Disease Spectrum

**DOI:** 10.64898/2026.05.25.26354067

**Authors:** Keshuo Lin, Perminder S. Sachdev, Jiyang Jiang, the Alzheimer’s Disease Neuroimaging Initiative

**Author notes:** Corresponding author: E.

## Abstract

Although the associations between cerebrovascular dysfunctions and Alzheimer’s disease are increasingly appreciated, the relationship of cerebral blood flow and white matter hyperintensities with tau and amyloid-β pathology remains unclear, particularly in the longitudinal context. This study investigated cross-sectional and longitudinal associations of cerebral blood flow and white matter hyperintensities with tau and amyloid-β pathology using multimodal imaging and blood biomarkers in 179 participants from the ADNI3 cohort. Participants underwent structural (T1-weighted, T2-weighted FLAIR) and arterial spin labelling perfusion MRI, tau and amyloid-β PET, and plasma assay tests for amyloid-β 42, amyloid-β 40, and phosphorylated tau-217. Tau from PET was negatively associated with cerebral blood flow both cross-sectionally and longitudinally in the posterior brain, independent of amyloid-β quantified from PET. Higher white matter hyperintensities volumes were associated with higher levels of tau and amyloid-β at baseline, but the associations were significantly attenuated after further adjusting for amyloid-β and tau, respectively. Plasma amyloid-β 42/40 ratio was negatively associated with white matter hyperintensity volumes both cross-sectionally and longitudinally. In conclusion, tau pathology showed spatially specific associations with cerebral hypoperfusion, independent of amyloid-β, particularly in posterior regions. The attenuation of associations of white matter hyperintensities with amyloid-β and tau after adjustment may reflect shared disease-related variance rather than distinct independent effects.

## Introduction

Alzheimer’s disease (AD) is a progressive neurodegenerative disorder characterized by Amyloid-β (Aβ) plaques, tau neurofibrillary tangles, and neurodegeneration, which are widely recognized as the hallmarks of the disease (DeTure & Dickson, 2019; Iaccarino et al., 2023; Jack et al., 2016). In recent years, vascular factors are increasingly implicated. Reduced cerebral blood flow (CBF) and increased white matter hyperintensity (WMH) volumes are common in the early stages of AD and may contribute to cognitive decline (Korte et al., 2020; Prins & Scheltens, 2015).

However, studies on the relationship of CBF and WMH with Aβ and tau have yielded inconsistent findings, with some showing significant associations of higher WMH (Cha et al., 2024; Graff-Radford et al., 2019) and lower CBF (Mattsson et al., 2014) with elevated levels of Aβ, but not tau, whereas others finding their associations with tau only (McAleese et al., 2015; Visser et al., 2023; Visser et al., 2020). There were also studies showing that WMH volumes (Garnier-Crussard et al., 2020) and CBF (Bangen et al., 2017) were not associated with Aβ or tau, and that lower CBF was associated with both higher Aβ and tau levels (Rubinski et al., 2021; Weigand et al., 2022). The longitudinal Korean Brain Aging Study demonstrated that baseline Aβ burden, but not tau, was associated with subsequent increases in WMH volume over a two-year period, particularly among females (Cha et al., 2024). A longitudinal study using Alzheimer’s Disease Neuroimaging Initiative (ADNI) data showed that increases in within-session intraindividual cognitive variability—defined as dispersion across neuropsychological tests administered at a single visit—from baseline to 12-month follow-up were significantly associated with concurrent reductions in CBF in the entorhinal and hippocampal regions, but only in individuals classified as Aβ-positive based on cerebrospinal fluid (CSF) pTau/Aβ42 ratios at the cut-off of 0.0198 (Holmqvist et al., 2022). The Baltimore Longitudinal Study of Aging (BLSA) and ADNI datasets showed that Aβ status was not associated with changes in CBF, glucose metabolism, or volumes of the inferior temporal gyrus, highlighting limited vascular impact of Aβ accumulation in this region (Bilgel et al., 2022). Notably, current literature investigating the relationships of cerebrovascular biomarkers with tau and Aβ usually focuses on AD-vulnerable regions. Whole brain patterns of these associations have not been well documented.

The Aβ cascade model of AD posits the primacy of Aβ deposition preceding tau-mediated neurofibrillary tangle formation (Hardy, 2017). However, emerging evidence suggests that tau pathology may develop independently of Aβ. A PET imaging study using ADNI data found that 46% of non-demented older adults exhibited tau accumulation in Braak stage I/II regions without cortical Aβ positivity (A−/T+). Individuals in the A−/T+ group showed worse cognitive performance compared to A−/T− individuals, and cognitive decline was associated with tau burden even in the absence of Aβ (Weigand et al., 2020). The underlying mechanisms driving WMH formation and CBF reduction may differ. Moreover, they appear at different stages during cerebrovascular dysfunction. The aetiology of WMH is diverse and may include neuroinflammation, cerebral microvascular damage, demyelination, and genetic influence (Sachdev et al., 2016; Wardlaw et al., 2015). The reduction of CBF, which is the primary cause for subsequent ischaemic vascular lesions, can stem from more systemic vascular factors such as blood pressure dysregulation (Korte et al., 2020). Given these distinct pathophysiological pathways, examining the associations of WMH and CBF with tau and Aβ will provide insights into how cerebrovascular dysfunction interacts with AD. As a result, the present study aims to examine both cross-sectional and longitudinal associations of CBF and WMH with Aβ and tau levels, using MRI and PET imaging data from the ADNI Phase 3 (ADNI3). We hypothesized that tau and Aβ pathology are differentially associated with vascular dysfunction, and the associations may be region specific.

## Methods

### Sample characteristics

To maintain the consistency of imaging data, the study included 179 participants from the ADNI3 cohort, of whom 122 were followed up for an average of 2.49 years. The dataset consists of 95 cognitively normal (CN), 63 with mild cognitive impairment (MCI), and 21 with AD. MRI data for all participants were acquired at 14 ADNI sites using the same scanner model, GE Discovery MR750w. Sample characteristics are summarized in Table 1.

**Table 1.** Sample Characteristics at Baseline. CBF - Cerebral Blood Flow; WMH - White Matter Hyperintensities; WM – white matter; GM – grey matter; WBWMH - Whole Brain WMH; PVWMH - Periventricular WMH; DWMH - Deep WMH; GM - Gray Matter; AD - Alzheimer’s Disease; CN - Cognitively Normal; MCI - Mild Cognitive Impairment; MoCA – Montreal Cognitive Assessment; MMSE – Mini-Mental State Examination; SUVR - Standardized Uptake Value Ratio; **^1^**meta temporal regions: bilateral entorhinal cortex, amygdala, fusiform, and inferior and middle temporal cortices. ^2^ a cut-off meta-temporal SUVR 1.27 used to determine tau positivity. ^3^ Aβ positivity was defined using established SUVR cut-offs: 1.08 for ^18^F-florbetaben and 1.11 for ^18^F-florbetapair. ^4^ pTau-217 data were available for a subset of 121 subjects in ADNI3 with 38 subjects (13 CN, 25 MCI) included in the current study.

|  |  | CN | MCI | AD | Total |
| --- | --- | --- | --- | --- | --- |
| N |  | 95 | 63 | 21 | 179 |
| Age (years) |  | 72.97 ± 7.01 | 74.76 ± 7.44 | 75.37 ± 5.92 | 73.88 ± 7.08 |
| Sex (male%) |  | 42.1% | 63.5% | 71.4% | 53.1% |
| Education (years) |  | 16.52 ± 2.63 | 16.68 ± 2.34 | 15.38 ± 2.71 | 16.44 ± 2.56 |
| MMSE |  | 29.34 ± 0.99 | 27.37 ± 2.92 | 23.57 ± 2.01 | 27.97 ± 2.71 |
| MoCA |  | 26.34 ± 2.91 | 23.05 ± 3.66 | 17.80 ± 4.27 | 24.20 ± 4.34 |
| PET | <sup>18</sup>F-florbetapir amyloid SUVR | 1.10 ± 0.20 | 1.19 ± 0.26 | 1.44 ± 0.00 | 1.14 ± 0.23 |
|  | <sup>18</sup>F-florbetaben amyloid SUVR | 1.12 ± 0.20 | 1.13 ± 0.25 | 1.60 ± 0.25 | 1.23 ± 0.30 |
|  | Meta temporal <sup>1</sup> tau SUVR | 1.21 ± 0.16 | 1.30 ± 0.31 | 1.73 ± 0.39 | 1.31 ± 0.30 |
|  | Tau<sup>+</sup> <sup>2</sup> | 20.0% | 30.2% | 81.0% | 30.7% |
|  | Aβ<sup>+</sup> <sup>3</sup> | 27.4% | 33.3% | 81.0% | 35.8% |
| WMH (mm<sup>3</sup>) | WBWMH | 7972.71 ± 20315.08 | 8332.98 ± 10616.92 | 8304.75 ± 7129.66 | 8138.46 ± 16215.69 |
|  | PVWMH | 4204.79 ± 8720.25 | 5191.29 ± 6531.47 | 5431.50 ± 5330.77 | 4695.91 ± 7647.80 |
|  | DWMH | 3503.82 ± 10789.12 | 3007.77 ± 4446.54 | 2783.25 ± 2351.29 | 3244.69 ± 8310.30 |
| CBF (mL/100g/min) | Total GM | 35.00 ± 8.54 | 30.72 ± 9.58 | 25.84 ± 9.76 | 32.42 ± 9.53 |
|  | Total WM | 29.16 ± 7.13 | 26.46 ± 7.60 | 22.18 ± 8.02 | 27.39 ± 7.71 |
|  | WMH | 26.04 ± 7.71 | 23.28 ± 8.51 | 18.98 ± 8.09 | 24.24 ± 8.33 |
|  | normal-appearing WM | 29.25 ± 7.11 | 26.55 ± 7.61 | 22.28 ± 8.03 | 27.48 ± 7.70 |
|  | Frontal | 31.04 ± 7.60 | 27.71 ± 8.22 | 23.91 ± 8.15 | 29.03 ± 8.20 |
|  | Temporal | 33.11 ± 7.69 | 29.60 ± 8.41 | 24.86 ± 7.98 | 30.90 ± 8.40 |
|  | Parietal | 34.45 ± 9.13 | 30.41 ± 10.02 | 24.57 ± 11.08 | 31.87 ± 10.17 |
|  | Occipital | 37.04 ± 11.56 | 30.58 ± 12.00 | 25.36 ± 13.79 | 33.40 ± 12.63 |
|  | Putamen | 32.04 ± 5.94 | 30.70 ± 7.49 | 28.75 ± 6.79 | 31.18 ± 6.67 |
|  | Accumbens | 29.93 ± 6.03 | 27.37 ± 8.41 | 25.26 ± 6.74 | 28.48 ± 7.19 |
|  | Hippocampus | 33.18 ± 6.29 | 30.36 ± 7.32 | 27.20 ± 6.67 | 31.48 ± 6.98 |
|  | Amygdala | 30.95 ± 6.13 | 28.76 ± 7.59 | 25.37 ± 6.49 | 29.52 ± 6.93 |
|  | Pallidum | 26.06 ± 4.97 | 25.47 ± 6.06 | 23.62 ± 5.10 | 25.57 ± 5.42 |
|  | Caudate | 26.00 ± 5.03 | 23.98 ± 6.98 | 21.26 ± 5.86 | 24.73 ± 6.05 |
|  | Thalamus | 32.97 ± 7.01 | 30.05 ± 8.74 | 25.71 ± 7.24 | 31.09 ± 8.01 |
|  | Brainstem | 27.06 ± 5.12 | 25.73 ± 6.28 | 22.71 ± 5.87 | 26.08 ± 5.78 |
| **Blood biomarkers** | pTau-217 (pg/mL) <sup>4</sup> | 1.94 ± 1.87 | 2.61 ± 2.36 | - | 2.23 ± 2.10 |
|  | Aβ42/Aβ40 ratio | 0.07 ± 0.03 | 0.06 ± 0.03 | 0.04 ± 0.02 | 0.07 ± 0.03 |
|  | Aβ42 | 26.81 ± 15.07 | 24.29 ± 10.85 | 12.61 ± 5.29 | 24.18 ± 13.56 |
|  | Aβ40 | 366.49 ± 100.91 | 370.97 ± 105.21 | 326.27 ± 95.96 | 362.99 ± 101.64 |

### MRI Acquisition

All participants had T1-weighted, T2-weighted FLAIR, and arterial spin labelling (ASL) perfusion MRI data acquired using 3T GE Discovery MR750w scanners following the ADNI3 imaging protocol. T1-weighted structural images were acquired using a 3D accelerated sagittal inversion recovery fast spoiled gradient echo sequence (repetition time (TR) = 10ms, echo time (TE) = minimum full, inversion time (TI) = 400ms, flip angle = 11°, bandwidth = 31.25kHz, field of view = 25.6cm, 1 mm slices, 256 × 256 matrix). T2-weighted FLAIR images were obtained using a 3D sagittal Cube sequence, with TR = 4800ms, TE = 119ms, TI = 1451ms, 1.2 mm slices, and matrix = 256 × 256. The 3D pseudo-continuous ASL sequence was acquired with labelling duration = 1.45s, post-labelling delay = 2.025s, spiral readout with 8 arms × 512 points, TR = 4897ms, TE = 10.7ms, flip = 111°, 4 mm slices. All images were visually inspected and met ADNI quality control standards. For T1-weighted MRI, image selection followed the Clinica ADNI-to-BIDS procedure, which uses MPRAGEMETA.csv, and MRIQUALITY.csv for scan selection and quality control. For ASL, scans were retained if they passed QC recorded in the ADNI UCSF ASL QC file (QCRating = “Pass”, Jack et al., 2024).

### WMH segmentation

WMH were segmented and quantified using the UBO Detector pipeline (Jiang et al., 2018). Briefly, FLAIR images were linearly registered to each subject’s T1-weighted image, followed by tissue segmentation on T1-weighted data. Deformation fields from native T1 space to Diffeomorphic Anatomical Registration Through Exponentiated Lie Algebra (DARTEL) space were then computed and applied to bring the FLAIR images to DARTEL space. After skull stripping, FSL FAST was used to segment the DARTEL-space FLAIR images into candidate WMH clusters. A k-nearest neighbour (k-NN) classifier with default settings (k = 5, probability threshold = 0.7) was applied to distinguish WMH from non-WMH clusters based on intensity, anatomical location, and cluster size features. Periventricular WMH (PVWMH) were defined as WMH voxels within 12 mm from the lateral ventricles, while voxels located ≥12 mm away were classified as deep WMH (DWMH). All resultant WMH masks were overlaid on corresponding FLAIR images for visual inspection. All results passed quality control.

### CBF quantification

The perfusion-weighted images (PWIs) were calibrated with the ventricular cerebrospinal fluid (CSF) signal from M0 scans using the BASIL pipeline in FSL (Chappell et al., 2023). Quantitative CBF was then estimated through kinetic model inversion, enabling conversion of the perfusion-weighted signal into absolute CBF values in mL/100 g/min. All ventricular masks underwent quality control, including visual inspection and manual editing, to ensure only retaining voxels confined to the ventricular regions.

When visualising PWIs, it is noticed that posterior brain regions had higher signal compared to anterior regions in some PWIs. Two additional methods were applied to reduce this posterior bias: 1) applying N4 bias field correction to PWIs before ventricular CSF calibration, and 2) using voxel-wise calibration option available in BASIL. CBF quantified by using these 2 methods were compared to the one calibrated with ventricular CSF through visual inspection, examining associations of CBF derived from each method with diagnosis, age and time. Results are described in Supplementary Text.

To quantify total grey matter and white matter CBF, the following workflow was implemented to register calibrated PWIs to the MNI152 standard space. First, T1-weighted images were skull-stripped using SynthStrip (Hoopes et al., 2022). Second, M0 images were rigidly registered to the T1 space using FSL FLIRT. Third, skull-stripped T1 images were affinely registered to the MNI152 template, followed by nonlinear registration using FSL FNIRT to compute warp fields. Finally, the calibrated PWI maps were warped to MNI152 space using the precomputed nonlinear warp and M0-to-T1 transformation matrix. All intermediate outputs—including registered T1 images, transformation matrices, and warped images—were saved for quality control and reproducibility. Grey and white matter masks were derived from tissue priors, with the grey matter threshold of 0.4 and white matter threshold of 0.5. To calculate CBF in the FreeSurfer DK atlas, the M0 image was co-registered to the native T1-weighted image. The resulting transformation was then applied to the CBF maps.

### Aβ and Tau PET Acquisition and Quantification

All Aβ and tau PET scans from the ADNI3 database were processed as previously described (Landau et al., 2025). Briefly, dynamic PET frames (5-min intervals) were motion-corrected by co-registering all frames to the first, averaged to produce a static image, and reoriented to a standardized 160 × 160 × 96 matrix (1.5 mm³ voxels) aligned along the anterior–posterior commissure axis. Standard uptake value ratios (SUVR) were calculated using cerebellar grey matter as the reference region.

In the current dataset, 71 participants with 119 Aβ PET scans were performed using ^18^F-florbetaben (FBB) tracer, and 86 individuals with 173 scans using ^18^F-florbetapir (FBP). Tau levels were measured by using ^18^F-flortaucipir ligand. Tau positivity was defined as a temporal meta-ROI flortaucipir SUVR ≥ 1.27, where the temporal meta-ROI included bilateral entorhinal cortex, amygdala, fusiform, and inferior and middle temporal cortices (Jack et al., 2017). Since two Aβ ligands (FBB and FBP) were used and they have different cut-offs for Aβ+, Aβ positivity was defined as a binary variable to maximize cross-sectional sample size, using established SUVR thresholds of 1.08 for FBB and 1.11 for FBP (Royse et al., 2021). For longitudinal analyses, only participants with FBB scans were included, and Aβ SUVR from FBB scans was analysed as a continuous variable.

### Blood biomarkers of Aβ and tau

All biofluid samples were processed and shipped following harmonized protocols across sites. Participants fasted for at least six hours (water only) prior to blood draws and lumbar puncture procedures (Weiner et al., 2017). Plasma levels of Aβ40, Aβ42, and phosphorylated tau-217 (pTau-217) were extracted from the FNIHBC_BLOOD_BIOMARKER_TRAJECTORIES file downloaded from ADNI website. All blood biomarkers included in the present study were measured using the Roche Elecsys platform. The Aβ42/Aβ40 ratio was used as the primary metric for blood-based Aβ pathology. In the present dataset, blood-based Aβ measures were available for 241 sessions, while pTau-217 measures were available for 90 sessions. Characteristics of blood biomarkers were summarised in Table 1.

### Neuropsychological tests

Mini-Mental State Examination, Montreal Cognitive Assessment, and Clinical Dementia Rating memory subscale scores were included in this study to examine associations with cognition.

### Vascular risk factors

Hypertension category was defined using systolic and diastolic blood pressure measurements as follows: normal blood pressure was defined as systolic blood pressure <120 mmHg and diastolic blood pressure <80 mmHg; boundary blood pressure was defined as systolic blood pressure 120–139 mmHg or diastolic blood pressure 80–89 mmHg; and hypertension was defined as systolic blood pressure ≥140 mmHg or diastolic blood pressure ≥90 mmHg. APOE ε4 status was coded according to the number of ε4 alleles, with 0 indicating no ε4 allele, 1 indicating one ε4 allele, and 2 indicating two ε4 alleles.

### Statistical Analysis

All statistical analyses were conducted using R (version 4.3.3). Linear regression models were used to investigate the associations of tau and Aβ with CBF and WMH. The primary models were adjusted for age, sex, APOE ε4 and hypertension. For CBF, the regression models were run for the whole brain and each region in the FreeSurfer DK atlas. For regional CBF analyses, false discovery rate (FDR) correction was applied to account for multiple comparisons across brain regions. Longitudinal associations of CBF and WMH with tau and Aβ burden were assessed using linear mixed-effects models with a random intercept for each participant. Random slopes were not included because the available longitudinal data were insufficient to reliably estimate subject-specific slopes, with the number of observations being less than or equal to the number of random effects when random-slope models were tested. CBF or WMH was used as the dependent variable, and tau or Aβ was included as the independent variable of interest. Models were constructed by separating within- and between-subject effects. Specifically, models included changes of imaging measures from baseline (within-subject effects) and baseline levels of these measures (between-subject effects). Time since baseline was also included in the linear mixed models.

## Results

The distribution of CBF, WMH, tau, and Aβ were summarised in Figure 1. Associations of CBF and WMH with cognitive performance, demographic factors, and Time were summarized in Supplementary Text.

**Figure 1.**
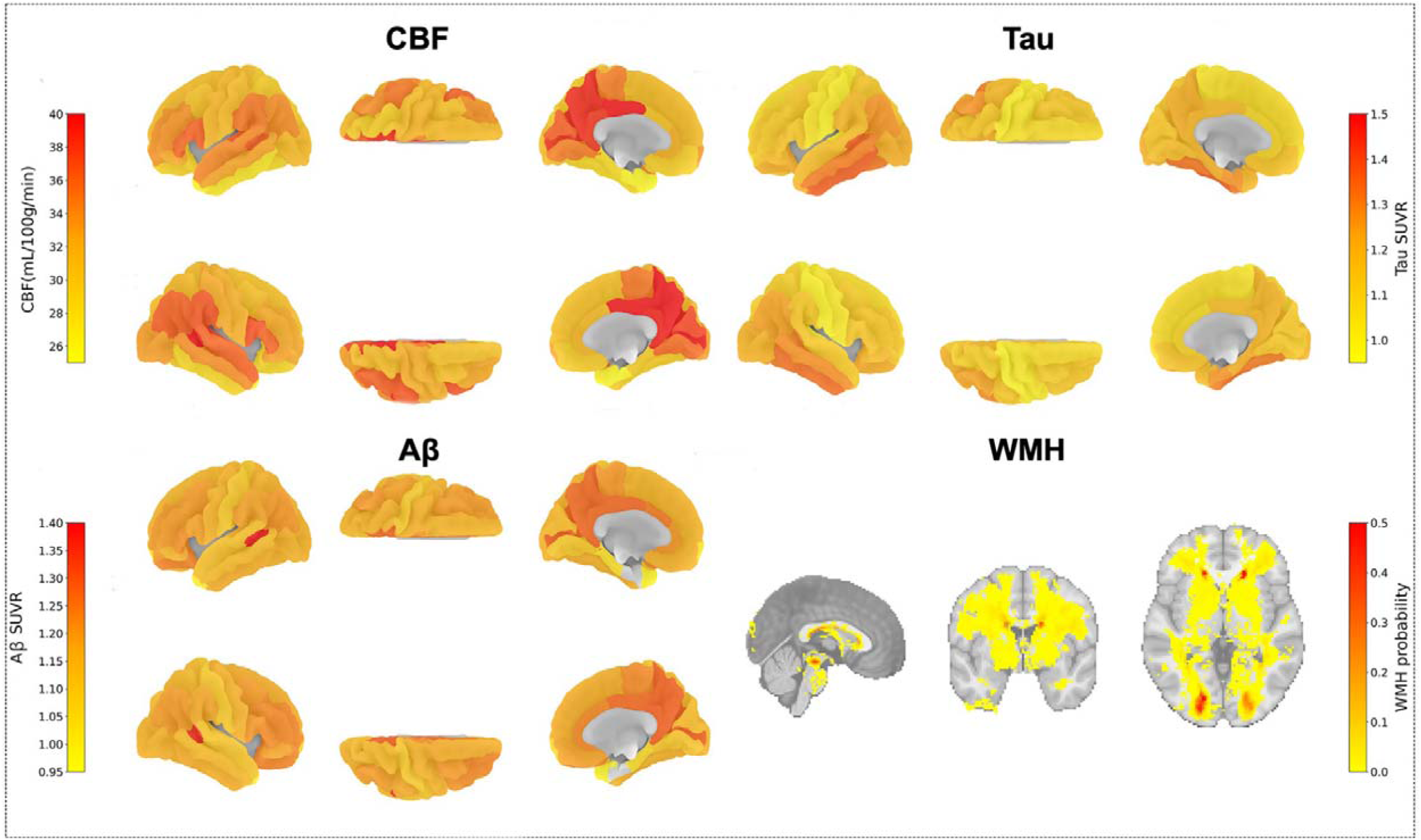
Spatial distribution of cerebral blood flow (CBF), white matter hyperintensities (WMH, Tau, and Amyloid-β (Aβ) SUVR at baseline.

### Baseline associations of CBF and WMH with tau and Aβ

Significantly negative associations between CBF and PET tau levels, after adjusting for age, sex, APOE ε4 and hypertension are summarised in Supplementary Table 1 and Figure 2a. Notably, associations were more significant in the posterior part of the brain, especially in the parietal and occipital areas. After FDR correction, significant associations remained in the caudal middle frontal, fusiform, inferior parietal, inferior temporal, isthmus cingulate, lateral occipital, middle temporal, precuneus, superior parietal, and supramarginal regions. Detailed regional results are presented in Supplementary Table 1. After further adjustment for Aβ, a similar spatial pattern was observed, but did not survive FDR correction (Supplementary Table 1 and Figure 2b). No significant associations were observed between Aβ and CBF in any examined regions (Supplementary Figure 1). Additional sensitivity analyses to examine whether partial volume effects in CBF maps and regional brain atrophy affected the findings are presented in the Supplementary Text and Supplementary Tables 3 and 5. Overall, these sensitivity analyses showed that the negative tau–CBF association pattern was largely preserved after accounting for partial volume effects and regional brain atrophy.

**Figure 2.**
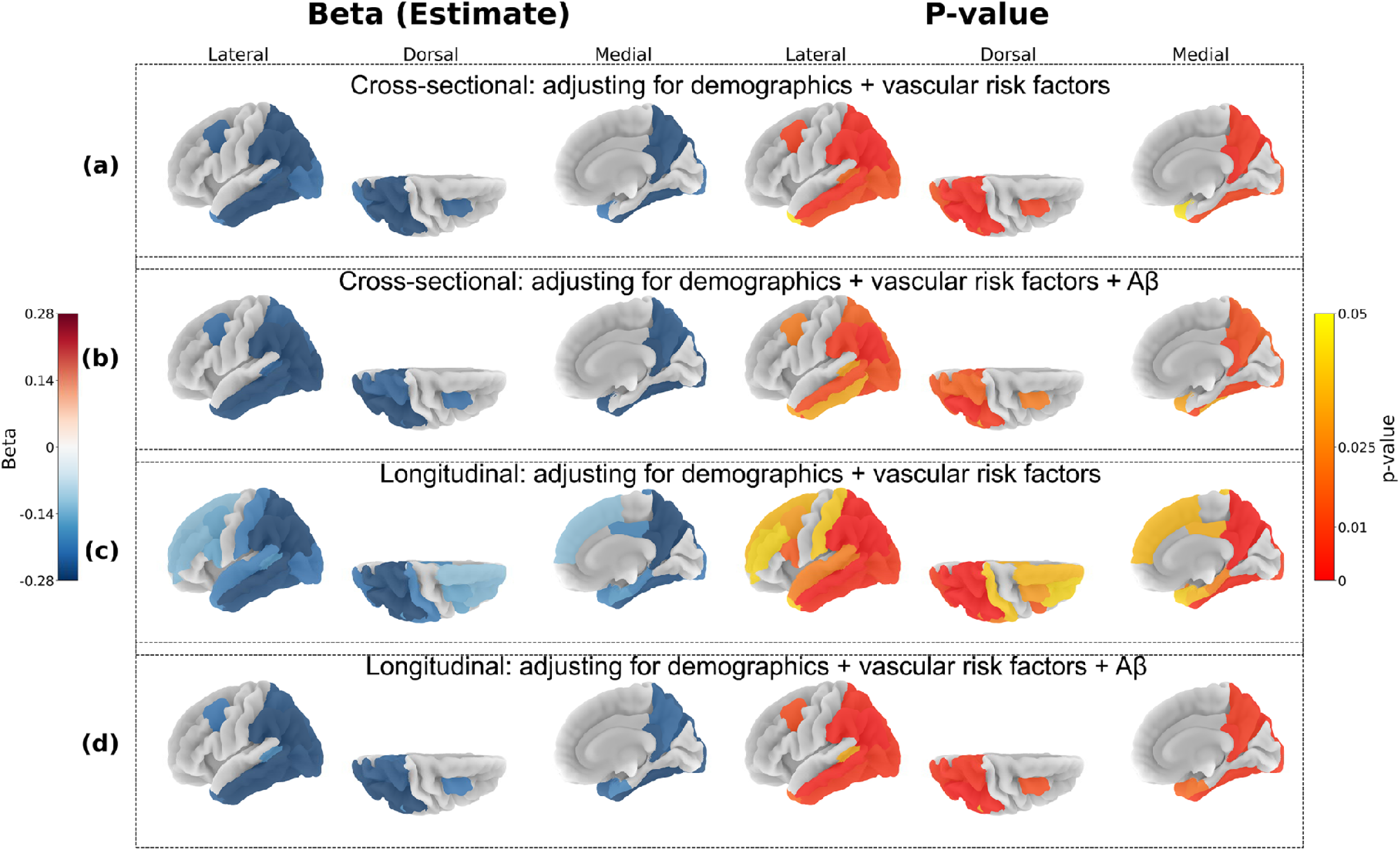
Associations between cerebral blood flow (CBF) and Tau levels. **(a)** Cross-sectional associations between CBF and tau at baseline, adjusting for age and sex. **(b)** Cross-sectional associations between CBF and tau after further adjusting for Aβ. **(c)** Within-subject effects in the longitudinal associations between changes in CBF and tau over time, after adjusting for baseline age, sex, hypertension and APOE ε4, using a linear mixed-effects model with random intercept. **(d)** within-subject effects in the longitudinal associations between changes in CBF and tau after further accounting for Aβ changes. For panels (a)–(d), only regions with significant associations (p[<[0.05) are shown. Left columns display maps of beta coefficients, where red indicates positive associations and blue indicates negative associations. Right columns show p-value maps, with red indicating stronger statistical significance (lower p-values).

The associations of WMH with tau and Aβ levels are summarized in Table 2. Higher tau levels were positively associated with higher WBWMH (β = 0.176, p = 0.042) and PVWMH (β = 0.181, p = 0.035) volumes, after adjusting for age, sex, APOE ε4 and hypertension. After further adjusting for Aβ status, these associations were no longer significant. Aβ was positively associated with WMH burden in the whole brain, and periventricular and deep WM regions (β = 0.490 – 0.563, p < 0.001). After adjusting for tau, these associations were significantly attenuated with only the association with DWMH remaining significant (β = 0.380, p = 0.035).

**Table 2.** Associations of white matter hyperintensity volumes with tau and Aβ. WMH – White Matter Hyperintensities; WBWMH – Whole Brain WMH; PVWMH – Periventricular WMH; DWMH – Deep WMH; FBB – ^18^F-Florbetaben; CI – Confidence Interval; CI lower – Lower bound of the 95% CI; CI upper – Upper bound of the 95% CI. *p*-values are annotated as follows: † *p* < 0.1; * *p* < 0.05; ** p < 0.001; *** p < 0.001.

| | Tau | | | Aβ | | |
| --- | --- | --- | --- | --- | --- | --- |
| | $\beta$ | CI lower | CI upper | $\beta$ | CI lower | CI upper |
| WBMWH | 0.176* | 0.006 | 0.345 | 0.534*** | 0.225 | 0.844 |
| PVWMH | 0.181* | 0.017 | 0.350 | 0.563*** | 0.256 | 0.871 |
| DWMH | 0.138† | -0.017 | 0.294 | 0.490*** | 0.208 | 0.772 |
| | Tau (additionally control for Aβ) | | | Aβ (additionally control for tau) | | |
| | $\beta$ | CI lower | CI upper | $\beta$ | CI lower | CI upper |
| WBMWH | 0.057 | -0.144 | 0.258 | 0.319 | -0.067 | 0.705 |
| PVWMH | 0.054 | -0.147 | 0.255 | 0.360 † | -0.026 | 0.746 |
| DWMH | 0.019 | -0.163 | 0.202 | 0.380 * | 0.027 | 0.732 |

### Longitudinal associations of CBF and WMH with tau and Aβ

Significantly negative associations between within-subject changes in tau and CBF over time were observed in widespread brain regions (Supplementary Table 2 and Figure 2c). After FDR correction, these associations remained significant in the fusiform, inferior parietal, inferior temporal, isthmus cingulate, lateral occipital, middle temporal, pars opercularis, precuneus, superior parietal, and supramarginal regions (Supplementary Table 2). After further adjustment for Aβ changes, nominally significant negative associations were observed in the Banks STS, caudal middle frontal, entorhinal, fusiform, inferior parietal, inferior temporal, isthmus cingulate, lateral occipital, middle temporal, occipital, precuneus, superior parietal, supramarginal, temporal pole, and frontal regions (Supplementary Table 2 and Figure 2d). After FDR correction, the Banks STS and frontal regions did not survive correction (Supplementary Table 2). Similar sensitivity analyses to investigate effects of partial volume effects and regional brain atrophy are presented in the Supplementary Text and Supplementary Tables 4 and 6. Overall, the sensitivity analyses were broadly consistent with the main longitudinal findings. However, after further adjustment for Aβ changes, some regions identified in the main analyses, such as lateral occipital, did not remain significant in the PVC-CBF or atrophy-adjusted models, whereas the banks of the superior temporal sulcus remained significant in both sensitivity analyses and the frontal composite remained significant only in the atrophy-adjusted model.

### Associations of blood-based biomarkers with WMH and CBF

Lower plasma Aβ42/40 ratio was significantly associated with higher volumes of WBWMH, PVWMH and DWMH (β = –0.260 - -0.398, p < 0.007). Longitudinal linear mixed-effects models showed that decreases in Aβ42/40 ratios were associated with greater increases in volumes of WBWMH, PVWMH, and DWMH (β = –0.278- -0.336, p < 0.003). No significant associations between pTau-217 and WMH was observed in any region, either cross-sectionally or longitudinally. No significant associations were found between blood biomarkers and total or regional GM CBF.

## Discussion

In the ADNI3 dataset, we examined associations of CBF and WMH with Aβ and tau across the AD spectrum. Tau was negatively associated with CBF both cross-sectionally and longitudinally, independent of Aβ, especially in the posterior brain. Greater WMH burden was also associated with greater cerebral tau and Aβ levels at baseline, but these associations were significantly attenuated after further adjusting for Aβ and tau, respectively. Additionally, WMH volumes were negatively associated with plasma Aβ42/40 ratios both cross-sectionally and longitudinally.

The findings indicate that higher tau burden is more consistently associated with lower CBF in posterior cortical regions than in anterior regions. According to Braak staging, the entorhinal cortex is one of the earliest regions affected by tau pathology in AD (Braak et al., 2006; Braak & Braak, 1995). A recent study mapped tau progression from the anterior temporal lobe to neocortical and posterior regions, including the parietal cortex (Berron et al., 2021). Studies have also shown that the regional distribution of tau in posterior brain areas strongly predicted future cortical atrophy (La Joie et al., 2020). In this context, the posterior predominance of tau–CBF associations observed in the present study may reflect spatial overlap between regions vulnerable to tau accumulation and regions showing reduced perfusion. However, the observational design does not establish whether tau contributes to CBF reduction, reduced CBF promotes tau accumulation, or both processes are related to broader disease severity.

The present findings show that associations between tau and CBF were observed after accounting for Aβ, whereas Aβ–CBF associations were not detected in this dataset. This pattern is consistent with previous work reporting associations between tau burden and CBF alterations that were not fully explained by Aβ status (Bilgel et al., 2021). Experimental studies have also suggested potential links between tau-related pathology and vascular or endothelial dysfunction (Hussong et al., 2023). Nevertheless, the current observational analyses cannot determine causal directionality or establish a direct mechanistic effect of tau on cerebral perfusion. The PET ligands used for Aβ in ADNI3 are highly selective for fibrillar Aβ and therefore primarily reflect neuritic plaque burden rather than diffuse plaques or vascular Aβ (Ikonomovic et al., 2008). Whether the absence of Aβ–CBF associations reflects biological specificity, measurement characteristics, or limited statistical sensitivity requires further investigation.

Baseline associations between WMH burden and both tau and Aβ were initially significant but became non-significant after adjusting for the other pathology. The attenuation of associations suggests that the associations between WMH and tau, and between WMH and Aβ, were largely from the same signal. Although WMH are often considered markers of cerebral small vessel diseases, accumulating evidence indicates that they can also arise from non-vascular mechanisms, such as AD-derived WMH (Garnier-Crussard et al., 2023). The disappearance of independent associations indicates that the influence may come from general neurodegenerative processes in AD, which are associated with both Aβ and tau levels.

In the present study, WMH burden demonstrated significant associations with both Aβ and tau pathology, whereas CBF alterations were selectively associated with tau pathology, and not with Aβ. These divergent patterns likely reflect differences in underlying biological mechanisms. WMH development has been linked to the accumulation of various plasma-derived proteins within the perivascular space, including fibrinogen, immunoglobulins, thrombomodulin, and potentially Aβ itself (Wardlaw et al., 2015). The infiltration of these proteins into the vessel walls may contribute to arteriolar wall thickening and subsequent white matter degeneration. Importantly, both Aβ deposition and WMH exhibit heterogeneous and spatially diffuse distributions across the brain, which may underlie their observed association. In contrast, CBF reductions tend to follow a more regionally specific and stereotyped pattern, closely mirroring the cortical propagation of tau pathology. This spatial concordance may explain the stronger and more consistent associations between CBF and tau.

The plasma Aβ42/40 ratio was negatively associated with WMH burden cross-sectionally and longitudinally, and PET-derived Aβ showed positive associations with WMH volumes at baseline. These findings are consistent with prior studies reporting that lower Aβ42/40 ratio (de Havenon et al., 2024; Hilal et al., 2017) and higher PET-derived Aβ levels (Graff-Radford et al., 2019; Lee et al., 2018) are associated with greater WMH burden and subclinical markers of cerebral small vessel disease. It is noteworthy that there are significant differences in the sensitivity between blood and PET methods to detect Aβ. A recent study demonstrated that plasma Aβ42/40, measured using a highly sensitive chemiluminescence immunoassay, identified Aβ positivity in 61.5% of participants with intermediate Aβ PET centiloid scores, while visual PET assessment classified only 20% of these participants as positive (Bun et al., 2023). Thus, blood based Aβ assays may detect subtle Aβ changes impacting vascular integrity at earlier stages, even before these alterations are visible on PET imaging.

Two key limitations warrant consideration. First, the relatively small sample sizes for FBB imaging and limited FBP data availability resulted in reduced statistical power for detecting longitudinal amyloid changes, potentially limiting our ability to capture clinically meaningful Aβ progression. Second, the inconsistent results observed between FBP and FBB measurements highlight the need for future mechanistic studies to elucidate the underlying causes of these discrepancies between different Aβ assessment methods.

## Conclusion

Tau pathology showed consistent associations with lower CBF, particularly in posterior cortical regions, after accounting for Aβ. WMH burden was associated with both tau and Aβ at baseline, although these associations were attenuated after mutual adjustment. Plasma Aβ42/40 ratio was inversely related to WMH burden. Overall, the findings demonstrate distinct association patterns of CBF and WMH with AD-related pathologies, while emphasizing that the causal direction and biological mechanisms underlying these relationships require further investigation.

## Data Availability

Data used in this study were obtained from the ADNI database (https://adni.loni.usc.edu). ADNI data are publicly available to qualified researchers upon registration and agreement to the ADNI Data Use Agreement. Data will be available beginning at the time of publication and for up to 2 years thereafter.

## Acknowledgment

The authors thank the ADNI participants for their invaluable contributions to research and the ADNI investigators for data collection and sharing.

## Funding Statement

Not applicable.

## Conflicts of interest

PSS reports receiving research funding from the National Health and Medical Research Council of Australia (APP1169489) and the National Institutes of Health, USA (grants 1RF1AG057531–01 and 2R01AG057531-02A1), with all payments made to his institution. He received an honorarium from Alkem Labs for a lecture as part of the Frontiers of Psychiatry 2023 seminar in Mumbai, India. He has served on the Medical Advisory Committees of Biogen Australia (2020–2021), Roche Australia (2022), Eli Lilly (2025) and Novo Nordisk (2025), with payment received for participation in meetings. He also holds unpaid leadership roles with the International Neuropsychiatric Association (Executive Board Member) and the World Psychiatric Association (Planning Committee Member). KL and JJ declare no conflicts of interest.

## Ethical approval

ADNI obtained ethics approval from the Institutional Review Board (IRB) of the University of Southern California and from the IRBs of all participating institutions. Written informed consent was obtained from all participants or their authorized representatives. The full list of participating IRBs and ethics committees is provided in the Supplementary Text.

## Consent to participate

All participants provided written informed consent, and data collection procedures adhered to established ethical standards with subsequent de-identification of all participant data.

## Consent for publication

Not applicable.

## Code availability

Not applicable.

## Author contribution statement

Keshuo Lin (KL): Data pre-processing; Formal analysis; Writing – original draft; Writing – review & editing.

Jiyang Jiang (JJ): Conceptualisation, data collection; Writing – review & editing.

Perminder S. Sachdev (PSS): Writing – review & editing.

All authors: Approved the final manuscript.

